# Cognitive Performance in Prodromal Neuronal α-Synuclein Disease

**DOI:** 10.64898/2026.11.05.26367361

**Authors:** Saar Anis, Eric Zimmerman, Scott A. Sperling, Jay L. Alberts, Daniel Weintraub, Tanya Simuni, Andrew Siderowf, Hubert H. Fernandez, the Parkinson’s Precision Medicine Initiative

**Author notes:** Corresponding author: Saar Anis, MD, Cleveland Clinic, 9500 Euclid Ave., Cleveland, OH 44195.

## Abstract

**Importance:** Cognitive abnormalities may emerge during prodromal neuronal α-synuclein disease (NSD), but their domain specificity and longitudinal course are uncertain.

**Objective:** To characterize baseline cognition and longitudinal change in prodromal NSD compared with healthy controls, NSD-negative genetic carriers, and sporadic Parkinson disease (PD).

**Design:** Longitudinal cohort study using Parkinson’s Precision Medicine Initiative (PPMI) data, with first visits from July 2010 through April 2026 and up to 4 years of follow-up.

**Setting:** Multicenter cohort.

**Participants:** Healthy controls (HC; n=244), *GBA1* or *LRRK2* nonmanifesting carriers without NSD (n=323), prodromal NSD participants recruited through hyposmia or isolated rapid eye movement sleep behavior disorder cohorts (n=1640), and sporadic PD (n=824).

**Exposures:** NSD status and clinical cohort.

**Main Outcomes and Measures:** Cognitive summary scores for verbal memory, executive function, processing speed, language, and visuospatial function, and PPMI-defined mild cognitive impairment (MCI). Models adjusted for age, sex, education, and APOE ε4 status.

**Results:** Among 3031 participants (mean [SD] age, 66.6 [7.0] years; 1322 female [43.6%]), prodromal NSD participants had lower baseline verbal-memory performance than HC (adjusted difference, −0.20 SD; 95% CI, −0.34 to −0.07; adjusted *P* = .007); NSD-negative genetic carriers did not differ from HC. No other cognitive domain was lower than HC at baseline or longitudinally. In the primary age-adjusted analysis, verbal-memory trajectory differed from HC over 4 years (*P* = .009), whereas in a smaller age-matched sensitivity subset through year 3, it did not (*P* = .62); the baseline difference persisted (adjusted difference, −0.28 SD; 95% CI, −0.46 to −0.10; *P* = .003). Adjusted MCI probability increased from 10.2% to 15.1% in prodromal NSD, compared with 8.4% to 5.0% in HC and 13.4% to 17.7% in PD. The increase was greater than in HC (ratio of annual odds ratios, 1.28; 95% CI, 1.04 to 1.58; adjusted *P* = .04) but not PD (*P* = .62).

**Conclusions and Relevance:** Prodromal NSD was associated with modestly lower verbal memory and increasing MCI probability relative to HC, without lower performance in other cognitive domains. The baseline verbal-memory difference was robust, whereas the longitudinal trajectory finding was not reproduced in the smaller age-matched sensitivity subset.

**Key Points:** *Question:* What cognitive differences characterize prodromal neuronal α-synuclein disease, and how do they change over time?

*Findings:* In this longitudinal cohort study of 3031 participants, prodromal neuronal α-synuclein disease was associated with lower verbal memory at baseline than healthy controls, while other cognitive domains were not lower. In the primary age-adjusted analysis, the verbal-memory trajectory differed from healthy controls over 4 years; in a smaller age-matched sensitivity subset through year 3, it did not differ significantly.

*Meaning:* Cognitive involvement in prodromal neuronal α-synuclein disease may be detectable as a modest verbal-memory signal, while longitudinal changes require confirmation.

## Introduction

Parkinson’s disease (PD) has traditionally been diagnosed after the emergence of clinically evident motor parkinsonism. The Neuronal Synuclein Disease Integrated Staging System (NSD-ISS) is centered on the presence of pathologic neuronal α-synuclein detected by a validated in vivo biomarker. (1) Staging subsequently incorporates evidence of dopaminergic dysfunction, assessed using dopamine transporter imaging, together with the degree of clinical and functional impairment. Stage 2 includes clinical signs or symptoms without functional impairment, with stage 2A occurring without and stage 2B with dopaminergic dysfunction. Stages 3 through 6 reflect progressively greater functional impairment, categorized as slight, mild, moderate, and severe, respectively. (1,2)

Cognition is a major non-motor feature of PD. Cognitive impairment is already common in newly diagnosed PD, and longitudinal cohorts show that mild cognitive impairment (MCI) can emerge or progress early in the disease course. (3–5) Cognitive changes have also been described during the prodromal period. In hyposmic at-risk individuals, hyposmia has been associated with subsequent decline in memory and language, while isolated REM sleep behavior disorder (iRBD) has been associated with faster cognitive decline and phenoconversion to clinically-defined PD or Dementia with Lewy Bodies (DLB). (6–8)

More recently, Parkinson’s Precision Medicine Initiative (PPMI) data demonstrated subtle, cross-sectional, global cognitive differences in NSD-ISS stage 2, particularly among participants with dopaminergic deficit or iRBD. (9) However, whether these differences are domain-specific and how they change longitudinally before PD or other neurodegenerative disease diagnosis remain unclear. It is also uncertain whether early cognitive differences are associated with biomarker-defined synuclein disease itself or are already present in individuals with genetic risk but no detectable synuclein pathology.

We therefore used PPMI data to compare five cognitive domains and MCI longitudinally across healthy controls (HC), *GBA1*/*LRRK2* non-manifesting carriers without NSD (NMC NSD-), prodromal participants with NSD (Prodromal NSD+), and sporadic PD (PD).

## Methods

PPMI datasets were accessed in May 2026. Participants were included from the PD, HC, and prodromal cohorts. (10) Prodromal participants were additionally classified by NSD status, defined by evidence of neuronal α-synuclein pathology on cerebrospinal fluid α-synuclein seed amplification assay (αSyn-SAA), yielding four groups. Prodromal NSD+ participants were restricted to those in the hyposmia or iRBD subgroups, whereas NMC NSD-participants were restricted to *GBA1* or *LRRK2* non-manifesting carriers without NSD, excluding those enrolled in the hyposmia or iRBD subgroups. PD participants were required to have NSD-positive sporadic PD without a pathogenic GBA1 or LRRK2 variant, and HC were required to be NSD-negative. To restrict analyses to age and education ranges represented across all groups, participants were limited to ages 50 to 85 years and at least 7 years of education.

Cognitive summary scores (CSS) were calculated for five domains. Letter-Number Sequencing (11), lexical fluency (FAS), semantic fluency (animal naming) (12), and the difference between Trails B and A scores were assigned to executive function. (13) The modified Boston Naming Test was assigned to language. (14) Hopkins Verbal Learning Test-Revised (HVLT-R) total recall and delayed recall were assigned to verbal memory. (15) The Symbol Digit Modalities Test (16) and Trails A were assigned to processing speed, and the Benton Judgment of Line Orientation (JLO) (17) to visuospatial function.

To calculate CSS, raw scores for each cognitive assessment were first z-scored in a similar manner to Weintraub et al. (18). Robust healthy controls were defined as HC participants with MoCA at baseline ≥27, MoCA at year 1 ≥26, and a decline in MOCA from baseline to year 1 of no more than 2 points. For each assessment, participant z-scores were defined as standard deviations away from the robust HC mean values. Participant z-scores beyond ±3 SD were winsorized to ±3 SD, and z-scores were redefined so positive values represented better performance. CSS were calculated as the average of their component z-scores. If any assessment required for a CSS was missing, that CSS was treated as missing. Because of skewness in the modified Boston Naming Test, values were reflected and Box-Cox transformed before z-scoring. MCI status was determined from the PPMI neuropsychological MCI classification and was not independently calculated in this analysis. The PPMI operational definition identifies MCI based on at least two scores more than 1.5 SD below the specified normative mean across the HVLT Total Recall, HVLT Recognition Discrimination, JLO, Letter-Number Sequencing, Semantic Fluency, and Symbol Digit Modalities assessments; investigator-assigned MCI or dementia status was not used. The MCI classification was analyzed separately from the continuous domain CSS; in particular, HVLT recognition discrimination contributed to MCI classification but not to the verbal-memory CSS.

Longitudinal CSS were modeled using generalized estimating equations (GEE). For each domain, groups with at least 50 participants at baseline were included, and follow-up continued while each included group had at least 50 participants with data at each included study year. Models included terms for baseline age, sex, education, APOE ε4 status, diagnostic group, time, and the group-by-time interaction. APOE ε4 was dichotomized as carrier or noncarrier; missing covariates were imputed using predictive mean matching for numeric variables and logistic regression for binary variables. Linear and categorical time specifications were compared using the quasi-likelihood under the independence model criterion, with the lower-QIC model selected (Supplementary Table S1). MCI was modeled using a binomial GEE, and the visuospatial model additionally adjusted for JLO version. Model results were summarized as estimated marginal means (group means after covariate adjustment). Baseline differences were assessed using Wald contrasts of marginal means; longitudinal differences were assessed using slope contrasts for linear-time models and joint Wald contrasts for categorical-time models. Prodromal NSD+ comparisons with HC and PD comprised the primary contrast family; remaining contrasts were secondary. Holm-Bonferroni correction was applied within each family.

As a supplemental exploratory analysis, differences were assessed between baseline NSD-ISS stages rather than between diagnostic groups. The cognitive domains and MCI were each modeled using the same framework as the group comparison replacing diagnostic group with baseline NSD-ISS stage. NSD+ participants were included, combining the PD and Prodromal NSD+ groups, and including stages with at least 50 participants at baseline. Because cognition contributes to NSD-ISS stage assignment, these stage-based cognitive analyses were considered descriptive and were not interpreted as independent evidence of an association between stage and cognition. Contrasts were not divided into primary and secondary families when adjusting for multiple comparisons.

Sensitivity analyses. The robustness of the verbal memory findings was examined in four analyses: handling of missing APOE ε4 (setting all missing values to carrier and, separately, to non-carrier); adjustment for CSF Alzheimer co-pathology using the p-tau181/Aβ42 ratio; splitting Prodromal NSD+ by clinical subtype (iRBD versus hyposmia); and age matching of Prodromal NSD+ participants to HC. The contribution of dopaminergic deficit to the stage gradient was examined by additionally adjusting the stage comparison for baseline minimum putamen dopamine transporter SPECT (DAT-SPECT) specific binding ratio (SBR).

All analyses were conducted using RStudio 2026.10.1, R version 4.6.1.

## Results

### Cohort characteristics

Table 1 summarizes the four groups. Among included participants, first PPMI visits occurred between July 2010 and April 2026. The Prodromal NSD+ group included 1,232 participants recruited through hyposmia (75.1%) and 408 through iRBD (24.9%). Prodromal NSD+ participants were older than the other groups, with a mean (SD) age of 68.6 (5.7) years, compared with 65.0 (7.8) years in HC, 62.0 (6.8) years in NMC NSD-, and 64.9 (7.7) years in PD. Motor burden was substantially lower in Prodromal NSD+ than in PD, with mean OFF-medication MDS-UPDRS III scores of 5.1 (6.0) and 22.5 (9.6), respectively. APOE ε4 status was missing in 48.6% of the Prodromal NSD+ group.

**Table 1.** Cohort characteristics at baseline.

|  | **HC<br>(N=244)** | **NMC NSD-<br>(N=323)** | **Prodromal NSD+<br>(N=1640)** | **PD<br>(N=824)** |
|---|---|---|---|---|
| **Subgroup** |  |  |  |  |
| Sporadic PD | 0 (0%) | 0 (0%) | 0 (0%) | 824 (100%) |
| Healthy Control | 244 (100%) | 0 (0%) | 0 (0%) | 0 (0%) |
| GBA | 0 (0%) | 152 (47.1%) | 0 (0%) | 0 (0%) |
| LRRK2 | 0 (0%) | 171 (52.9%) | 0 (0%) | 0 (0%) |
| Hyposmia | 0 (0%) | 0 (0%) | 1232 (75.1%) | 0 (0%) |
| iRBD | 0 (0%) | 0 (0%) | 408 (24.9%) | 0 (0%) |
| **Age,** years | 65.0 (7.78) | 62.0 (6.75) | 68.6 (5.70) | 64.9 (7.68) |
| **Sex** |  |  |  |  |
| Female | 100 (41.0%) | 196 (60.7%) | 739 (45.1%) | 287 (34.8%) |
| Male | 144 (59.0%) | 127 (39.3%) | 901 (54.9%) | 537 (65.2%) |
| **Race** |  |  |  |  |
| White | 216 (88.5%) | 313 (96.9%) | 1556 (94.9%) | 763 (92.6%) |
| Black | 11 (4.5%) | 0 (0%) | 16 (1.0%) | 12 (1.5%) |
| Asian | 5 (2.0%) | 0 (0%) | 17 (1.0%) | 15 (1.8%) |
| Other | 12 (4.9%) | 8 (2.5%) | 38 (2.3%) | 30 (3.6%) |
| Missing | 0 (0%) | 2 (0.6%) | 13 (0.8%) | 4 (0.5%) |
| **Education,** years | 16.4 (2.71) | 16.9 (2.74) | 16.3 (2.61) | 16.1 (2.68) |
| **APOE ε4** |  |  |  |  |
| 0 alleles | 158 (64.8%) | 266 (82.4%) | 592 (36.1%) | 596 (72.3%) |
| 1 allele | 47 (19.3%) | 56 (17.3%) | 233 (14.2%) | 171 (20.8%) |
| 2 alleles | 6 (2.5%) | 0 (0%) | 18 (1.1%) | 11 (1.3%) |
| Missing | 33 (13.5%) | 1 (0.3%) | 797 (48.6%) | 46 (5.6%) |
| **Disease duration**, years | - | - | - | 0.75 (0.61) |
| **MDS-UPDRS I,** score | 3.22 (3.13) | 5.18 (4.33) | 6.77 (4.91) | 6.20 (4.37) |
| **MDS-UPDRS II,** score | 0.44 (0.97) | 1.03 (1.95) | 2.06 (3.20) | 6.05 (4.25) |
| **MDS-UPDRS III (OFF),** score | 1.53 (2.28) | 2.62 (3.67) | 5.06 (6.04) | 22.5 (9.58) |

Data are mean (SD) or n (%). NMC NSD-, GBA/LRRK2 non-manifesting carriers without NSD; Prodromal NSD+, prodromal participants with NSD recruited through hyposmia or iRBD.
Abbreviations: HC, healthy controls; NSD, neuronal synuclein disease; PD, Parkinson's disease; MDS-UPDRS I-III, Movement Disorder Society Unified Parkinson's Disease Rating Scale Parts I-III, OFF-medication state.

Verbal memory, visuospatial function, and MCI were analyzed through year 4; processing speed, executive function, and language through year 2, excluding NMC NSD-because only five carriers had baseline data. Fewer Prodromal NSD+ participants reached later years because of more recent enrollment (Figure 1; Supplementary Table S2).

**Figure 1.**
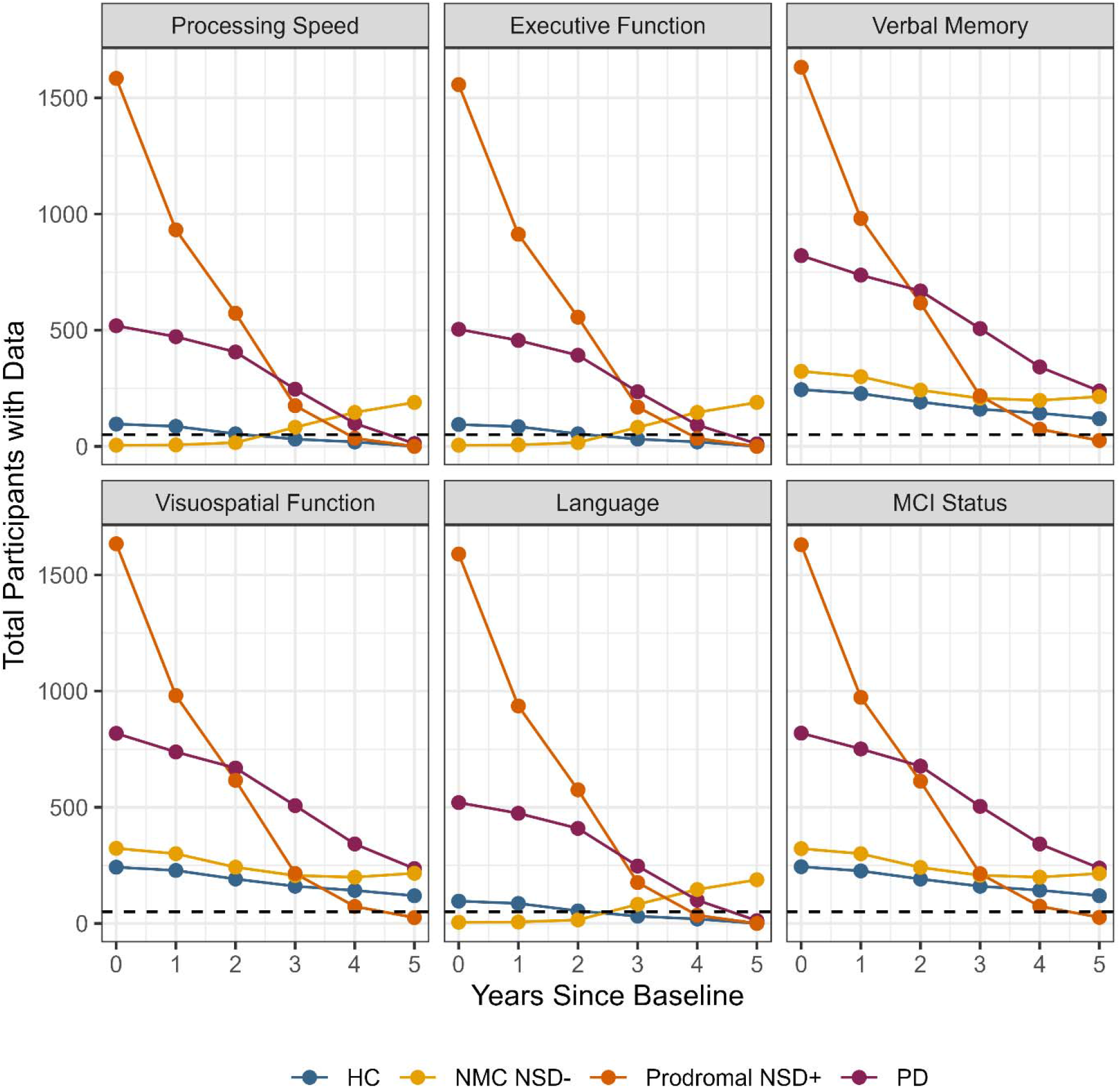
Participant availability by outcome and study year. Lines show the number of participants with non-missing data for each outcome by group and study year. The dashed horizontal line marks 50 participants, the prespecified minimum used to determine outcome-specific follow-up windows. NMC NSD-is absent from the processing speed, executive function, and language models because only five carriers had these measures at baseline. The shorter follow-up in Prodromal NSD+ primarily reflects more recent enrollment.

### Verbal Memory

Due to a better QIC, verbal memory was modeled using a categorical fit of time. At baseline, Prodromal NSD+ participants performed 0.20 SD below HC (95% CI, −0.34 to −0.07; adjusted *P* = .007) and 0.19 SD below NMC NSD-(95% CI, −0.31 to −0.07; adjusted *P* = .003). Prodromal NSD+ performed 0.08 SD better than PD at baseline (95% CI, −0.01 to 0.17; adjusted *P* = .08), although this difference was not statistically significant. NMC NSD-did not differ from HC (0.02 SD; *P* = .84).

The Prodromal NSD+ group showed worsening verbal-memory performance during the first 2 years. The overall trajectory differed significantly from HC (*P* = .009), for which performance was relatively constant over time, but not significantly different from PD (*P* = .18), for which there was similarly worse performance after baseline. Performance in the NMC NSD-group remained approximately unchanged from baseline to year 2 but showed a later drop in performance in years 3 and 4. The overall trajectory of the NMC NSD-group differed significantly from HC (*P* = .006) and Prodromal NSD+ (*P* < .001). Complete pairwise verbal memory contrasts are provided in Supplementary Table S3. Adjusted trajectories across the cognitive domains and MCI are shown in Figure 2. Differences between the Prodromal NSD+ group and the HC remained robust following the sensitivity analysis for missing APOE ε4 (Supplementary Table S4). When missing APOE ε4 were coded as carriers, the difference between the Prodromal NSD+ and PD group became significantly different (*P* = .022).

**Figure 2.**
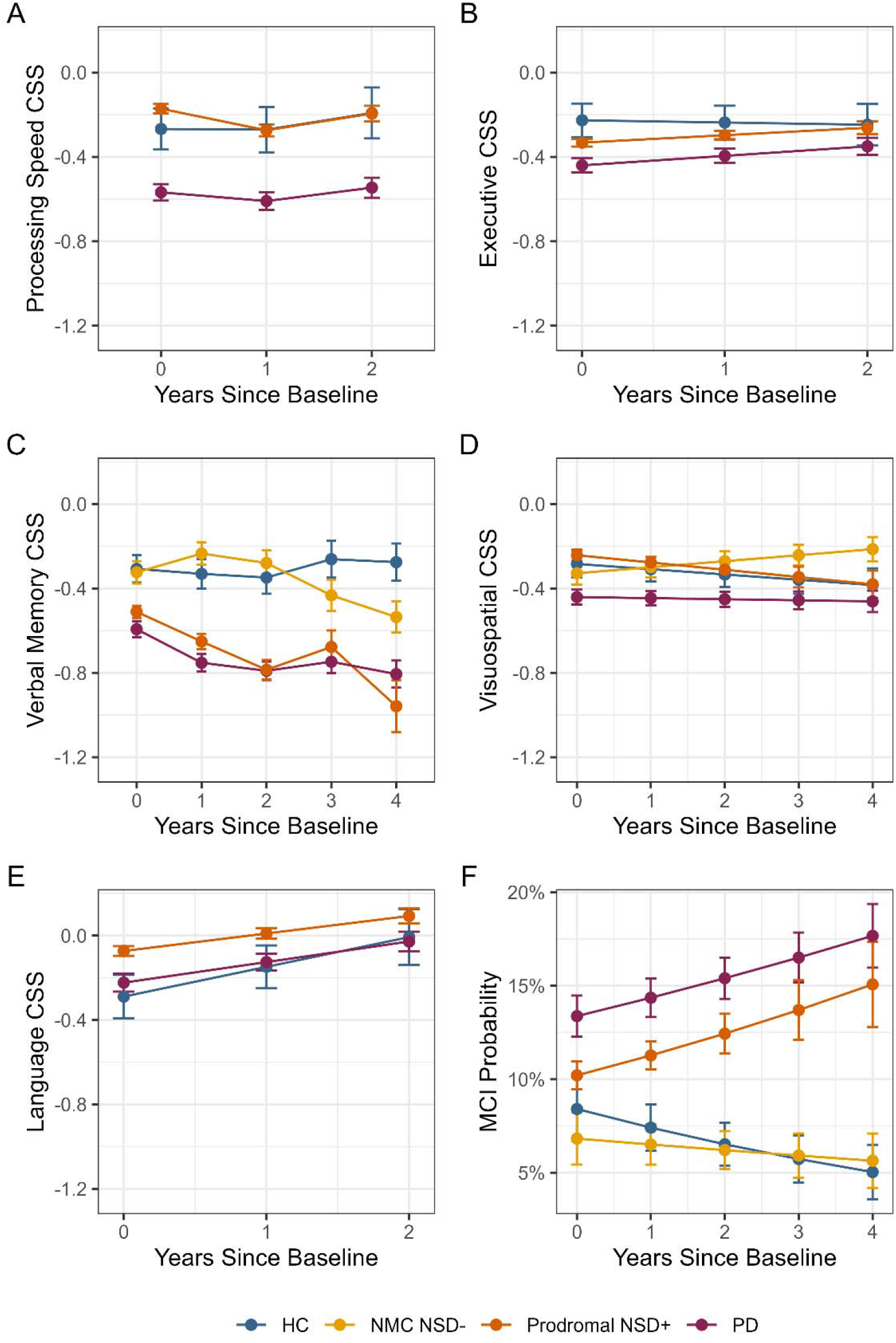
Cognitive and MCI trajectories by group, presented as estimated marginal means and standard errors from the corresponding GEE models. MCI marginal means (Panel F) are back-transformed to probabilities of MCI. NMC NSD-is shown where adequate cognitive data were available. Different follow-up windows reflect outcome-specific data availability. HC, healthy controls; NMC, non-manifesting carriers; NSD, neuronal synuclein disease; PD, Parkinson’s disease.

At baseline, the verbal memory difference from HC was numerically larger in the iRBD subgroup (0.31 SD, *P* = .002) than in the hyposmia subgroup (0.17 SD, *P* = .06), although the direct iRBD-hyposmia contrast did not reach multiplicity-adjusted significance (0.14 SD, *P* = .06). With follow-up restricted to year 3 because of smaller subgroup samples, the hyposmia trajectory differed from HC (*P* = .04), showing a decline in the first two years, whereas the iRBD trajectory did not differ (*P* = .60); the subgroup trajectories did not differ significantly from one another (*P* = .41). These analyses were exploratory (Figure 3; Supplementary Table S5).

**Figure 3.**
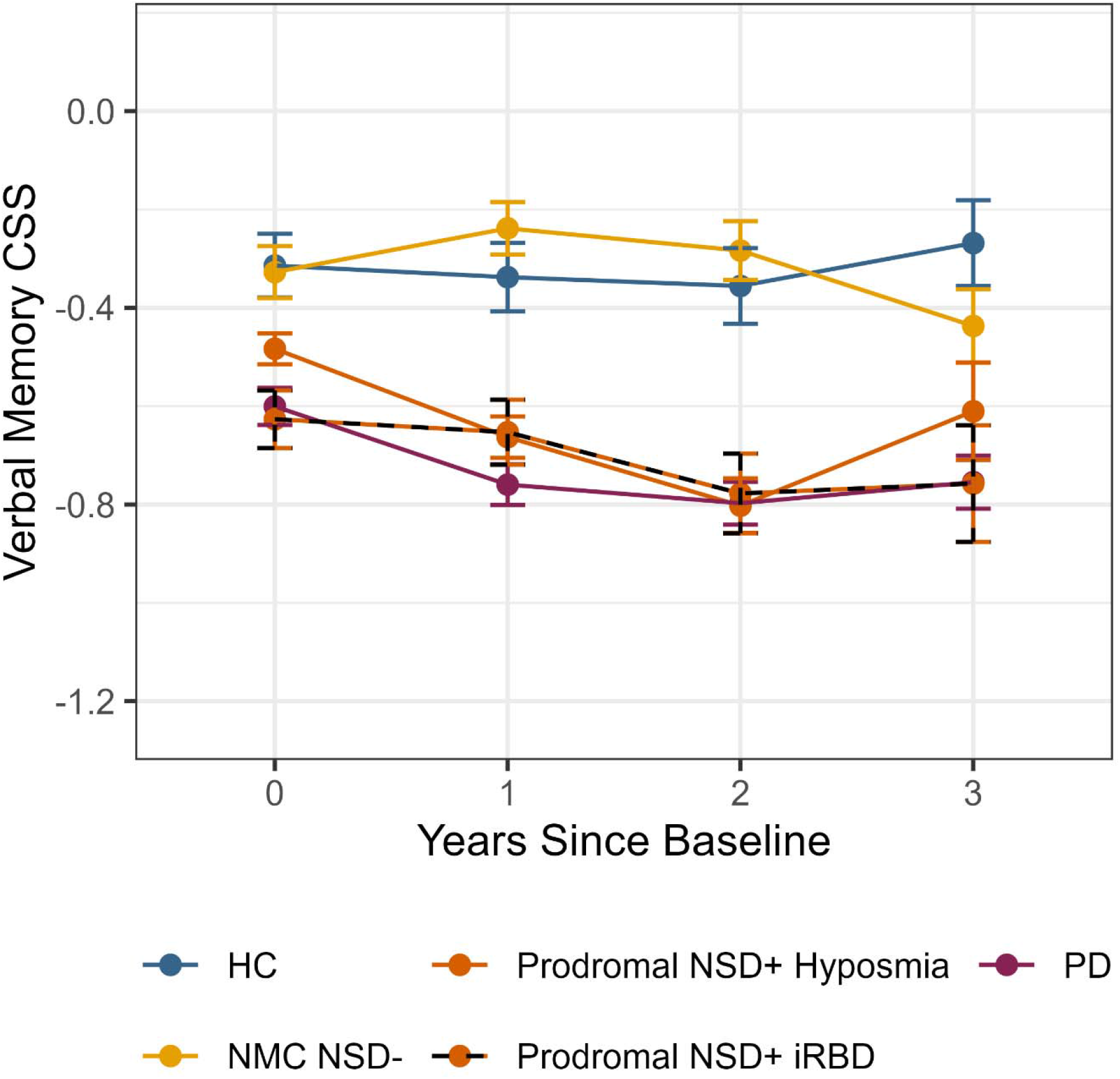
Verbal-memory trajectories after splitting Prodromal NSD+ by hyposmia and iRBD, presented as estimated marginal means and standard errors. Adjusted verbal-memory estimates are shown through year 3. The iRBD subgroup had the larger baseline difference from HC, while the hyposmia subgroup showed a significant longitudinal difference from HC. Direct iRBD-hyposmia baseline and longitudinal contrasts were not statistically significant after multiplicity correction.

### Other Cognitive Domains

At baseline, Prodromal NSD+ did not differ from HC in processing speed (*P* = .33), executive function (*P* = .19), or visuospatial ability (*P* = .54). Language performance was modestly higher in Prodromal NSD+ than in HC (0.22 SD, *P* = .04) and PD (0.15 SD, *P* = .003). PD showed lower performance than Prodromal NSD+ in all four non-memory domains (all *P* < .009) (Supplementary Table S6A). Longitudinal trends in processing speed, executive function, visuospatial ability, and language did not differ between Prodromal NSD+ and HC (all *P* ≥ .45) (Supplementary Table S6B).

### CSF Co-Pathology Sensitivity Analysis

Because a verbal memory-predominant pattern in an older cohort may reflect concomitant Alzheimer pathology, we repeated the baseline memory analysis in participants with available CSF p-tau181/Aβ42. The subset included 133 HC, 482 Prodromal NSD+ participants, and 557 participants with PD. Mean p-tau181/Aβ42 ratios were 0.05, 0.07, and 0.05, respectively. After adjustment for the CSF ratio, verbal memory remained significantly better in HC than Prodromal NSD+ (0.34 SD; 95% CI, 0.15-0.53; adjusted *P* < .001). HC also remained better than PD (0.36 SD, *P* < .001), whereas PD and Prodromal NSD+ did not differ (*P* = .73).

### Age-Matched Sensitivity Analysis

As an additional sensitivity analysis to address residual concern regarding the baseline age imbalance, 234 HC were matched to 234 Prodromal NSD+ participants on age; mean age was 65.5 years in both matched groups. In the matched sample, verbal memory remained lower in Prodromal NSD+ at baseline by 0.28 SD (95% CI, 0.10 to 0.46; *P* = .003). With follow-up modeled through year 3, the verbal memory trajectory did not differ significantly between Prodromal NSD+ and HC (*P* = .62) (Figure 4; Supplementary Table S7).

**Figure 4.**
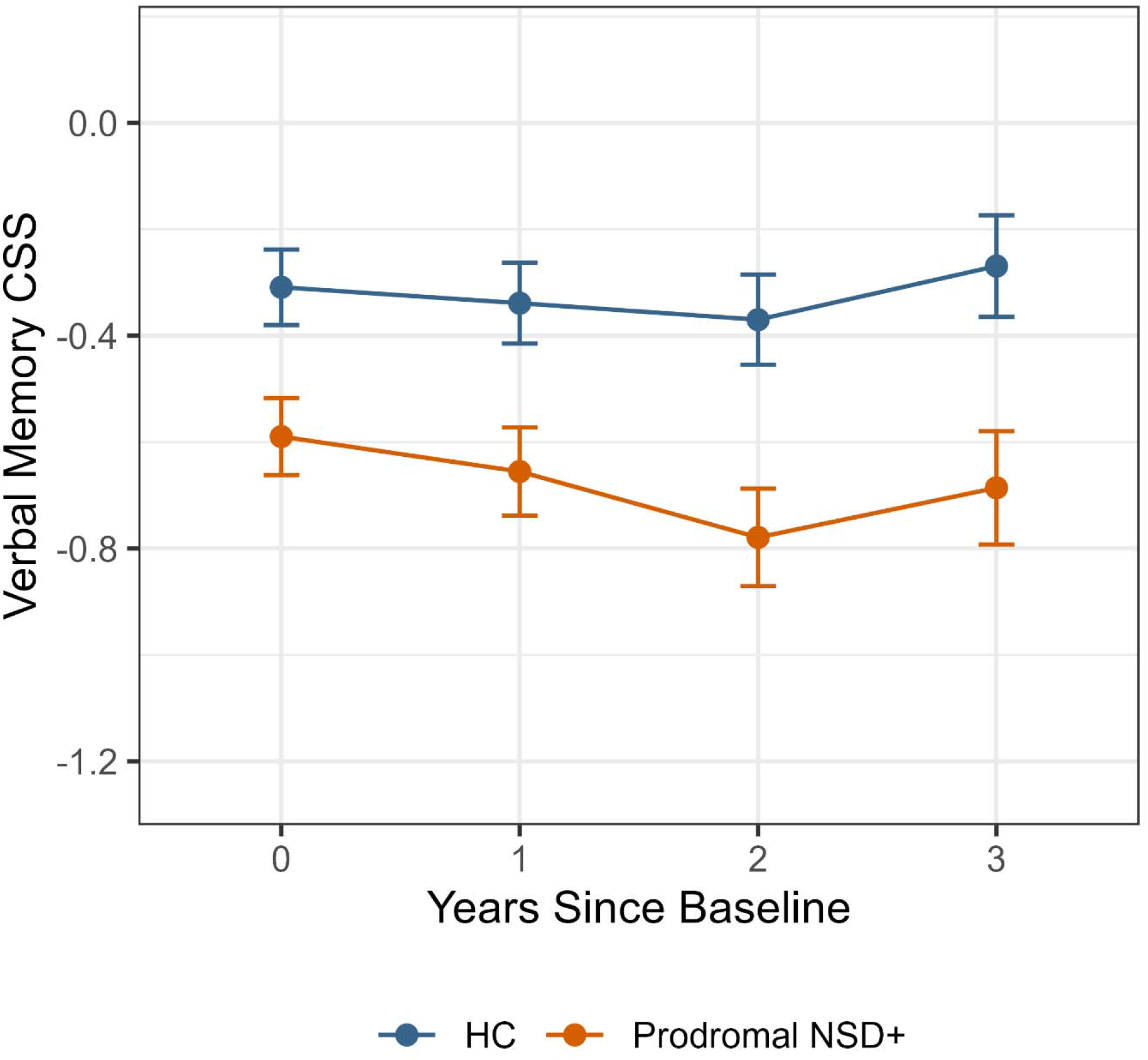
Age-matched sensitivity analysis of verbal memory, presented as estimated marginal means and standard errors. Adjusted verbal-memory estimates are shown for 234 HC and 234 Prodromal NSD+ participants matched on age. Prodromal NSD+ remained lower than HC at baseline (0.28 SD, *P* = .003), whereas the longitudinal trajectories through year 3 did not differ significantly (*P* = .62).

### Mild Cognitive Impairment

Adjusted MCI probability increased from 10.2% at baseline to 15.1% at year 4 in Prodromal NSD+. Corresponding adjusted probabilities were 13.4% to 17.7% in PD, 8.4% to 5.0% in HC, and 6.8% to 5.6% in NMC NSD-. At baseline, PD had higher adjusted odds of MCI than Prodromal NSD+ (odds ratio, 1.36; adjusted *P* = .02), whereas HC did not significantly differ from Prodromal NSD+ (odds ratio, 0.81; adjusted *P* = .36). Over 4 years, the increase in odds of MCI was greater in Prodromal NSD+ than in HC (ratio of annual odds ratios, 1.28; 95% CI, 1.04 to 1.58; adjusted *P* = .04), whereas longitudinal change did not differ between Prodromal NSD+ and PD (*P* = .62) (Supplementary Table S8).

### Exploratory NSD-ISS Stage Analyses

Exploratory longitudinal analyses by baseline NSD-ISS stage showed evidence of stage-specific trajectories for processing speed (stage-by-time *P* = .02), visuospatial function (*P* = .04), and language (*P* = .02), but not verbal memory (*P* = .19) or executive function (*P* = .19). MCI trends did not significantly differ between stages (*P* = .39). In the baseline verbal memory analysis, selected stage contrasts showed worse baseline performance at more advanced stages, but the specific contrasts changed after additional adjustment for baseline minimum putamen DAT-SPECT SBR, with no pairwise contrast significant in both models (Supplementary Tables S9-S10 and Supplementary Figure S1). Because cognition contributes to NSD-ISS stage assignment, all stage analyses were considered descriptive and were not used to support the primary conclusions.

## Discussion

In this longitudinal PPMI study, prodromal NSD was associated with a modest baseline verbal-memory difference relative to HC that was absent in NSD-negative genetic carriers. The baseline difference persisted after adjustment for CSF Alzheimer co-pathology and age matching. Verbal-memory and MCI trajectories differed from HC in the primary age-adjusted analyses, although the longitudinal verbal-memory trajectory difference was not reproduced in the smaller age-matched sensitivity subset. These findings suggest that cognitive involvement may be detectable before conventional PD diagnosis, with the strongest evidence for a modest baseline verbal-memory difference.

The prominence of verbal memory is consistent with prior evidence that memory impairment can be an important component of cognitive dysfunction in PD (19), while prior prodromal studies have also implicated memory and other cognitive domains. (20) In PD, HVLT-R performance has been shown to include distinct encoding-and retrieval-impaired profiles, with recognition performance helping differentiate these patterns. (21) In newly diagnosed, drug-naïve PD, verbal learning performance has also been linked to executive dysfunction and reduced semantic clustering, further supporting a contribution from frontal-subcortical processes. (22) Because recognition was not included in the verbal-memory CSS and narrative memory was unavailable, these findings should not be interpreted as an isolated medial-temporal amnestic syndrome. The absence of parallel processing-speed or executive deficits warrants replication.

The iRBD and hyposmia split further suggested heterogeneity within Prodromal NSD+. The baseline difference from HC was numerically larger in iRBD (0.31 SD) than hyposmia (0.17 SD), although the direct subgroup contrast did not reach multiplicity-adjusted significance. This direction is consistent with prior PPMI work showing greater cognitive impairment among participants with early NSD and hyposmia when iRBD was also present, including cognitive differences associated with iRBD even in the absence of dopaminergic dysfunction. (9) The longitudinal subgroup pattern was not parallel, however: hyposmia, but not iRBD, differed from HC over the available 3-year follow-up, and the two subgroup trajectories did not differ significantly. Separately, NMC NSD-participants were similar to HC in baseline verbal memory and performed better than Prodromal NSD+. Their trajectory itself differed from HC, so this comparator supports baseline specificity but not longitudinal stability.

The longitudinal MCI finding provides a complementary signal of cognitive involvement in Prodromal NSD+. Adjusted MCI probability increased from 10.2% at baseline to 15.1% at year 4, whereas it remained low in HC, and the longitudinal pattern differed significantly from HC but not PD. This is notable because the continuous domain analyses identified verbal memory as the only domain with lower performance at baseline, whereas the MCI definition includes HVLT recognition discrimination, which was not part of the verbal-memory CSS, as well as measures of visuospatial function, working memory, semantic fluency, and processing speed. The increasing probability of MCI may therefore capture broader or more heterogeneous cognitive changes that are not evident as group-level differences in individual domain CSSs. The absence of a significant longitudinal difference from early PD further raises the possibility that clinically-relevant cognitive change is already emerging before motor-driven PD diagnosis, although this should not be interpreted as evidence of equivalent trajectories.

### Limitations

This study has several limitations. Prodromal NSD+ participants were older than the other groups. Although primary models adjusted for age and analyses were restricted to age ranges represented across all groups, an additional age-matched sensitivity analysis confirmed the baseline verbal-memory difference but not the longitudinal trajectory difference. Follow-up was shorter in the more recently-enrolled Prodromal NSD+ cohort, although verbal memory data remained available for 617 participants at year 2 and 217 at year 3, with 74 participants contributing at year 4. The verbal-memory CSS included only HVLT-R total learning and delayed recall. However, word list learning measures, including the HVLT-R, are sensitive to cognitive change in PD and are well suited to longitudinal assessment. (23) Recognition contributed to MCI classification but not the verbal-memory CSS, and story or narrative memory was unavailable. In NMC NSD-, complete data were available only for verbal memory and visuospatial ability. The CSF analysis used a single p-tau181/Aβ42 ratio in a subset, and iRBD/hyposmia subgroup samples were smaller and clinically heterogeneous. Finally, NSD-ISS stage analyses were partially circular because cognition contributes to stage assignment and were treated as exploratory.

## Conclusion

Prodromal NSD+ participants had a small baseline verbal-memory difference from HC that was absent in NMC NSD- and persisted after CSF adjustment and age matching. In the primary age-adjusted analysis, the verbal-memory trajectory and adjusted MCI probability differed from HC, although the trajectory difference was not reproduced in the smaller age-matched sensitivity subset. Together, these findings support verbal memory and increasing MCI probability as early cognitive signals in NSD, while the longitudinal memory finding requires confirmation.

## Data Availability

Data used in the preparation of this article were obtained on 2026-05-11 from the Parkinson's Precision Medicine Initiative (PPMI) database (https://www.ppmi-info.org/access-data-specimens/download-data), RRID:SCR_006431. For up-to-date information on the study, visit http://www.ppmi-info.org.
This analysis used Tier 3 PPMI data, including DaTscan and αSyn-SAA results for prodromal participants, obtained upon request after approval by the PPMI Data Access Committee.

## Acknowledgments

Data used in the preparation of this article were obtained on 2026-05-11 from the Parkinson’s Precision Medicine Initiative (PPMI) database (https://www.ppmi-info.org/access-data-specimens/download-data), RRID:SCR_006431. For up-to-date information on the study, visit http://www.ppmi-info.org.

This analysis used Tier 3 PPMI data, including DaTscan and αSyn-SAA results for prodromal participants, obtained upon request after approval by the PPMI Data Access Committee.

Statistical analysis codes used to perform the analyses in this article are shared on GitHub https://github.com/CCF-Eric-Zimmerman/PPMI-Prodromal-Neuronal-Alpha-synuclein-Project and were given a persistent identifier via Zenodo 10.5281/zenodo.23979761.

Protocol information for The Parkinson’s Precision Medicine Initiative (PPMI) Clinical - Establishing a Deeply Phenotyped PD Cohort can be found on protocols.io or by following this link: https://dx.doi.org/10.17504/protocols.io.n92ldmw6ol5b/v2.

PPMI – a public-private partnership – is funded by the Michael J. Fox Foundation for Parkinson’s Research and funding partners, including AbbVie, Alamar Biosciences, Aligning Science Across Parkinson’s (ASAP), Arrowhead Pharma, Arvinas, AskBio, BIAL, BioArctic, Biohaven, BlueRock Therapeutics, Bristol Myers Squibb, Calico Labs, Capsida Biotherapeutics, Critical Path Institute, DaCapo Brainscience, Denali, Edmond J. Safra Foundation, Eli Lilly, Gain Therapeutics, GE Healthcare, Genentech, GSK, Insitro, Johnson & Johnson Innovative Medicine, Lundbeck, Merck, Neumora, Neuron23, Novartis, Olink, Regeneron, Roche, Sanofi, Tenvie, UCB, Vanqua Bio, Voyager Therapeutics, The Weston Family Foundation.

Use of Artificial Intelligence: ChatGPT (GPT-5.6 Sol; OpenAI) was used in October 2026 to assist with manuscript language editing, organization, condensation, and refinement of the title and abstract for clarity and journal style. It was not used to conduct the primary statistical analyses or generate the underlying study data. All AI-assisted content was reviewed and verified by the authors, who take full responsibility for the accuracy and integrity of the manuscript.

## Notes

### Competing Interest Statement

The authors have declared no competing interest.

### Clinical Protocols

https://dx.doi.org/10.17504/protocols.io.n92ldmw6ol5b/v2

### Author Declarations

Data used in the preparation of this article were obtained on 2026-05-11 from the Parkinson's Precision Medicine Initiative (PPMI) database (https://www.ppmi-info.org/access-data-specimens/download-data), RRID:SCR_006431. For up-to-date information on the study, visit http://www.ppmi-info.org. This analysis used Tier 3 PPMI data, including DaTscan and αSyn-SAA results for prodromal participants, obtained upon request after approval by the PPMI Data Access Committee.

