## Supplementary Material for "Cognitive Performance in Prodromal Neuronal α-Synuclein Disease"

This supplement presents outcome-specific data availability, complete contrast results, sensitivity analyses, and exploratory analyses that are not fully tabulated in the main manuscript. Group definitions and statistical methods are provided in the main manuscript. Unless otherwise indicated, p values use the multiple-comparison procedures specified in the main Methods.

### Supplementary Table S1. Longitudinal GEE model specification by cognitive domain

| **Cognitive summary score** | **Linear QIC** | **Categorical QIC** | **Selected time specification** |
| --- | --- | --- | --- |
| Processing speed | 3320 | 3316 | Categorical |
| Executive function | 2275 | 2277 | Linear |
| Verbal memory | 9819 | 9809 | Categorical |
| Visuospatial function | 9191 | 9196 | Linear |
| Language | 3812 | 3813 | Linear |

*For each cognitive domain, linear and categorical representations of time were compared using the quasi-likelihood under the independence model criterion (QIC), with the lower-QIC time specification selected. Abbreviations: GEE, generalized estimating equations; QIC, quasi-likelihood under the independence model criterion.*

### Supplementary Table S2. Participant availability by outcome, group, and study year

| **Cognitive summary score** | **Group** | **Year 0** | **Year 1** | **Year 2** | **Year 3** | **Year 4** | **Year 5** |
| --- | --- | --- | --- | --- | --- | --- | --- |
| Processing Speed | HC | 96 | 86 | 54 | 31 | 19 | 0 |
| Processing Speed | NMC NSD- | 5 | 6 | 16 | 82 | 146 | 189 |
| Processing Speed | Prodromal NSD+ | 1,584 | 932 | 573 | 175 | 35 | 0 |
| Processing Speed | PD | 519 | 472 | 406 | 246 | 99 | 12 |
| Executive function | HC | 94 | 85 | 54 | 31 | 19 | 0 |
| Executive function | NMC NSD- | 5 | 6 | 16 | 82 | 146 | 189 |
| Executive function | Prodromal NSD+ | 1,557 | 913 | 556 | 169 | 34 | 0 |
| Executive function | PD | 504 | 456 | 392 | 235 | 92 | 11 |
| Verbal memory | HC | 244 | 227 | 191 | 160 | 143 | 119 |
| Verbal memory | NMC NSD- | 323 | 300 | 242 | 207 | 198 | 214 |
| Verbal memory | Prodromal NSD+ | 1,632 | 981 | 617 | 217 | 74 | 25 |
| Verbal memory | PD | 821 | 737 | 669 | 507 | 342 | 238 |
| Visuospatial function | HC | 242 | 228 | 191 | 160 | 142 | 119 |
| Visuospatial function | NMC NSD- | 323 | 300 | 242 | 206 | 199 | 216 |
| Visuospatial function | Prodromal NSD+ | 1,634 | 981 | 616 | 215 | 73 | 25 |
| Visuospatial function | PD | 818 | 738 | 669 | 507 | 342 | 236 |
| Language | HC | 96 | 86 | 54 | 31 | 19 | 0 |
| Language | NMC NSD- | 5 | 6 | 15 | 82 | 146 | 188 |
| Language | Prodromal NSD+ | 1,590 | 936 | 575 | 176 | 35 | 0 |
| Language | PD | 520 | 474 | 409 | 247 | 100 | 12 |
| MCI status | HC | 244 | 226 | 191 | 160 | 143 | 119 |
| MCI status | NMC NSD- | 322 | 300 | 241 | 207 | 199 | 215 |
| MCI status | Prodromal NSD+ | 1,630 | 973 | 612 | 214 | 74 | 26 |
| MCI status | PD | 819 | 751 | 677 | 504 | 342 | 238 |

*Counts reflect non-missing outcome data entering the outcome-specific longitudinal analyses. The shorter follow-up in Prodromal NSD+ primarily reflects more recent enrollment. Participant counts are plotted in Figure 1 in the main manuscript. Abbreviations: HC, healthy controls; MCI, mild cognitive impairment; NMC NSD−, non-manifesting carriers without neuronal α-synuclein disease; PD, Parkinson’s disease; Prodromal NSD+, prodromal neuronal α-synuclein disease.*

### Supplementary Table S3. Complete pairwise verbal-memory comparisons

| **Contrast** | **Baseline estimate** | **Baseline *P*** | **Longitudinal joint F** | **Longitudinal *P*** |
| --- | --- | --- | --- | --- |
| HC vs Prodromal NSD+ | 0.204 | 0.0069 | 3.78 | 0.009 |
| PD vs Prodromal NSD+ | -0.082 | 0.075 | 1.56 | 0.182 |
| HC vs PD | 0.286 | 0.0003 | 2.25 | 0.061 |
| HC vs NMC NSD- | 0.016 | 0.842 | 4.03 | 0.0057 |
| PD vs NMC NSD- | -0.270 | <0.0001 | 6.64 | <0.0001 |
| NMC NSD- vs Prodromal NSD+ | 0.187 | 0.0034 | 6.96 | <0.0001 |

*For baseline estimates, the direction shown in the second column defines the sign of the estimate. Longitudinal p values are from joint Wald contrasts for the categorical-time memory model. P values use the primary/secondary contrast families described in the main Methods. Abbreviations: HC, healthy controls; NMC NSD−, non-manifesting carriers without neuronal α-synuclein disease; PD, Parkinson’s disease; Prodromal NSD+, prodromal neuronal α-synuclein disease; F, F statistic.*

### Supplementary Table S4. Sensitivity analysis for missing APOE ε4

| **Sensitivity assumption** | **HC vs Prodromal NSD+ estimate** | ***P*** | **PD vs Prodromal NSD+ estimate** | ***P*** | **NMC NSD- vs Prodromal NSD+ estimate** | ***P*** | **HC vs Prodromal NSD+ longitudinal *P*** |
| --- | --- | --- | --- | --- | --- | --- | --- |
| Missing APOE ε4 coded as non-carrier | 0.223 | 0.00282 | -0.063 | 0.174 | 0.209 | 0.00090 | 0.0182 |
| Missing APOE ε4 coded as carrier | 0.180 | 0.0218 | -0.115 | 0.0218 | 0.151 | 0.0298 | 0.00296 |

*The baseline HC-Prodromal NSD+ difference and longitudinal contrast remained statistically significant under both extreme coding assumptions for missing APOE ε4. Because outcome availability varies over follow-up, recoding missing APOE ε4 can affect the fitted longitudinal estimates despite APOE ε4 itself being time invariant.* *Abbreviations: APOE ε4, apolipoprotein E ε4 allele; HC, healthy controls; NMC NSD−, non-manifesting carriers without neuronal α-synuclein disease; PD, Parkinson’s disease; Prodromal NSD+, prodromal neuronal α-synuclein disease.*

### Supplementary Table S5. Pairwise verbal-memory comparisons after splitting Prodromal NSD+ by hyposmia and iRBD

| **Contrast** | **Baseline estimate** | **Baseline *P*** | **Longitudinal joint F** | **Longitudinal *P*** |
| --- | --- | --- | --- | --- |
| HC vs Prodromal NSD+ Hyposmia | 0.169 | 0.0626 | 3.993 | 0.0374 |
| HC vs Prodromal NSD+ iRBD | 0.312 | 0.00149 | 0.797 | 0.603 |
| PD vs Prodromal NSD+ Hyposmia | -0.117 | 0.0626 | 1.544 | 0.603 |
| PD vs Prodromal NSD+ iRBD | 0.026 | 0.701 | 1.287 | 0.603 |
| Prodromal NSD+ Hyposmia vs iRBD | 0.143 | 0.0626 | 2.057 | 0.414 |
| HC vs PD | 0.286 | 0.000353 | 2.555 | 0.119 |
| HC vs NMC NSD- | 0.013 | 0.871 | 2.780 | 0.119 |
| PD vs NMC NSD- | -0.273 | 0.000060 | 7.894 | 0.000118 |
| NMC NSD- vs Prodromal NSD+ Hyposmia | 0.156 | 0.0205 | 10.782 | 0.00000228 |
| NMC NSD- vs Prodromal NSD+ iRBD | 0.299 | 0.000492 | 1.747 | 0.155 |

*The subgroup model was restricted to year 3 because of smaller subgroup samples. The baseline difference from HC was larger in magnitude for iRBD (0.31 SD) than hyposmia (0.17 SD), but the direct iRBD-hyposmia contrast was not significant after correction (P = 0.0626). The hyposmia trajectory differed from HC (P = 0.0374), whereas the iRBD trajectory did not (P = 0.603). These subgroup analyses are exploratory. The primary contrast family used for the multiple comparison correction is defined as each of hyposmia and iRBD against HC, PD, and each other. Abbreviations: HC, healthy controls; iRBD, isolated rapid eye movement sleep behavior disorder; NMC NSD−, non-manifesting carriers without neuronal α-synuclein disease; PD, Parkinson’s disease; Prodromal NSD+, prodromal neuronal α-synuclein disease; SD, standard deviation; F, F statistic.*

### Supplementary Table S6A. Primary baseline contrasts by cognitive summary score

| **Cognitive summary score** | **Preferred fit** | **HC vs Prodromal NSD+ estimate** | ***P*** | **PD vs Prodromal NSD+ estimate** | ***P*** |
| --- | --- | --- | --- | --- | --- |
| Processing speed | Categorical | -0.096 | 0.328 | -0.396 | <0.0001 |
| Executive function | Linear | 0.106 | 0.189 | -0.108 | 0.0081 |
| Verbal memory | Categorical | 0.204 | 0.0069 | -0.082 | 0.075 |
| Visuospatial function | Linear | -0.041 | 0.537 | -0.198 | <0.0001 |
| Language | Linear | -0.216 | 0.039 | -0.150 | 0.0028 |

*Estimates are adjusted differences in CSS z-score. Positive HC - Prodromal NSD+ values indicate better performance in HC; negative PD - Prodromal NSD+ values indicate lower performance in PD. P values are Holm-adjusted within the primary contrast family. Abbreviations: CSS, cognitive summary score; HC, healthy controls; PD, Parkinson’s disease; Prodromal NSD+, prodromal neuronal α-synuclein disease.*

### Supplementary Table S6B. Primary longitudinal contrasts by cognitive summary score

| **Cognitive summary score** | **Preferred fit** | **HC vs Prodromal NSD+ longitudinal test** | ***P*** | **PD vs Prodromal NSD+ longitudinal test** | ***P*** |
| --- | --- | --- | --- | --- | --- |
| Processing speed | Categorical | Joint F=1.49 | 0.451 | Joint F=1.48 | 0.451 |
| Executive function | Linear | Slope difference=-0.046 SD/year | 0.585 | Slope difference=0.010 SD/year | 0.629 |
| Verbal memory | Categorical | Joint F=3.78 | 0.009 | Joint F=1.56 | 0.182 |
| Visuospatial function | Linear | Slope difference=0.009 SD/year | 0.723 | Slope difference=0.029 SD/year | 0.339 |
| Language | Linear | Slope difference=0.058 SD/year | 0.715 | Slope difference=0.015 SD/year | 0.715 |

*For domains modeled with categorical time, longitudinal differences were tested using joint Wald contrasts across follow-up and are summarized by the joint F statistic. For domains modeled with linear time, estimates are differences in annual slopes. P values are Holm-adjusted within the primary contrast family. Abbreviations: CSS, cognitive summary score; HC, healthy controls; PD, Parkinson’s disease; Prodromal NSD+, prodromal neuronal α-synuclein disease; SD, standard deviation; F, F statistic.*

### Supplementary Table S7. Age-matched sensitivity analysis of verbal memory

| **Measure** | **HC** | **Prodromal NSD+** | **Comparison/result** |
| --- | --- | --- | --- |
| Matched participants, baseline n | 234 | 234 | 1:1 age-matched sample |
| Age, mean (SD), years | 65.5 (7.53) | 65.5 (7.51) |  |
| Female, n (%) | 95 (40.6%) | 90 (38.5%) |  |
| Education, mean (SD), years | 16.5 (2.71) | 16.5 (2.65) |  |
| APOE ε4 missing, n (%) | 32 (13.7%) | 51 (21.8%) |  |
| Participants with memory data, year 1 | 217 | 175 |  |
| Participants with memory data, year 2 | 182 | 158 |  |
| Participants with memory data, year 3 | 152 | 119 |  |
| Baseline verbal-memory contrast |  |  | HC vs Prodromal NSD+ = 0.281 SD; *P =* 0.00263 |
| Longitudinal joint contrast, years 0-3 |  |  | F=0.60; *P =* 0.615 |

*Matching was performed between HC and Prodromal NSD+ using age. Year 4 was not included in the matched longitudinal model because only 41 matched Prodromal NSD+ participants had year-4 memory data. The matched analysis confirms the baseline difference but does not reproduce the longitudinal trajectory difference observed in the full cohort. Abbreviations: APOE ε4, apolipoprotein E ε4 allele; HC, healthy controls; n, number of participants; Prodromal NSD+, prodromal neuronal α-synuclein disease; SD, standard deviation; F, F statistic.*

### Supplementary Table S8. Complete pairwise MCI comparisons

| **Contrast** | **Baseline odds ratio** | **Baseline *P*** | **Trend contrast estimate** | **Longitudinal *P*** |
| --- | --- | --- | --- | --- |
| HC vs Prodromal NSD+ | 0.81 | 0.355 | -0.249 | 0.0397 |
| PD vs Prodromal NSD+ | 1.36 | 0.020 | -0.029 | 0.622 |
| HC vs PD | 0.59 | 0.084 | -0.219 | 0.114 |
| HC vs NMC NSD- | 1.25 | 0.467 | -0.086 | 0.504 |
| PD vs NMC NSD- | 2.11 | 0.0062 | 0.133 | 0.319 |
| NMC NSD- vs Prodromal NSD+ | 0.65 | 0.123 | -0.162 | 0.319 |

*Baseline odds ratios compare MCI odds in the first group with the second. Longitudinal contrasts are on the logit scale. P values are multiplicity-adjusted according to the primary and secondary contrast families in the main Methods. Abbreviations: HC, healthy controls; MCI, mild cognitive impairment; NMC NSD−, non-manifesting carriers without neuronal α-synuclein disease; PD, Parkinson’s disease; Prodromal NSD+, prodromal neuronal α-synuclein disease.*

### Supplementary Table S9. Exploratory baseline verbal-memory contrasts by NSD-ISS stage, before and after additional adjustment for baseline minimum putamen DAT-SPECT SBR

| **NSD-ISS stage contrast** | **Estimate** | ***P*** | **Estimate additionally adjusted for baseline minimum putamen SBR** | **SBR-adjusted *P*** |
| --- | --- | --- | --- | --- |
| 2a vs 2b | 0.113 | 0.143 | -0.010 | 1 |
| 2a vs 3 | 0.199 | 0.00252 | 0.060 | 1 |
| 2a vs 4 | 0.292 | 0.00320 | 0.260 | 0.115 |
| 2b vs 3 | 0.086 | 0.258 | 0.080 | 0.746 |
| 2b vs 4 | 0.179 | 0.143 | 0.270 | 0.0288 |
| 3 vs 4 | 0.093 | 0.289 | 0.190 | 0.157 |

Stage analyses are descriptive because cognition contributes to NSD-ISS stage assignment. Stage models adjusted for age, sex, education, and APOE ε4. The SBR-adjusted model additionally included baseline minimum putamen dopamine transporter SPECT (DAT-SPECT) specific binding ratio (SBR), defined as the lower SBR from the left and right putamen. The specific contrasts reaching statistical significance changed after this additional adjustment; no single pairwise stage contrast was statistically significant in both models. Accordingly, these results are not used to support the primary conclusions. Abbreviations: APOE ε4, apolipoprotein E ε4 allele; DAT-SPECT, dopamine transporter single-photon emission computed tomography; NSD-ISS, neuronal α-synuclein disease integrated staging system; SBR, specific binding ratio.

### Supplementary Table S10. Exploratory longitudinal cognitive and MCI trajectory analyses by baseline NSD-ISS stage

| **Outcome** | **Longitudinal stage test** | **Result** | **Holm-adjusted pairwise trajectory contrasts** |
| --- | --- | --- | --- |
| Processing speed | Stage by categorical time omnibus | F(6, 4286)=2.5; *P =* 0.021 | Stage 3 vs 4, *P =* 0.025; stage 2a vs 4, *P =* 0.0283 |
| Executive function | Stage by numeric time omnibus | F(3, 4145)=1.6; *P =* 0.193 | None significant |
| Verbal memory | Stage by categorical time omnibus | F(9, 5947)=1.4; *P =* 0.193 | None significant |
| Visuospatial function | Stage by numeric time omnibus | F(3, 5948)=2.7; *P =* 0.0437 | Stage 2a vs 3, *P =* 0.045 |
| Language | Stage by numeric time omnibus | F(3, 4313)=3.4; *P =* 0.018 | Stage 2a vs 4, *P =* 0.014 |
| MCI status | Stage by numeric time omnibus | F(3, 5967)=1.0, *P =* 0.392 | None significant |

*Cognitive-domain rows summarize GEE models using the selected time specification corresponding to the trajectories displayed in Supplementary Figure S1. For MCI, longitudinal stage differences were evaluated using pairwise contrasts of linear trends on the logit scale; all six pairwise comparisons were non-significant after Holm adjustment. These stage analyses are exploratory because cognition contributes to NSD-ISS stage assignment. Abbreviations: F, F statistic; GEE, generalized estimating equations; MCI, mild cognitive impairment; NSD-ISS, neuronal α-synuclein disease integrated staging system.*

**Supplementary Figure S1. Exploratory cognitive and MCI trajectories by baseline NSD-ISS stage**

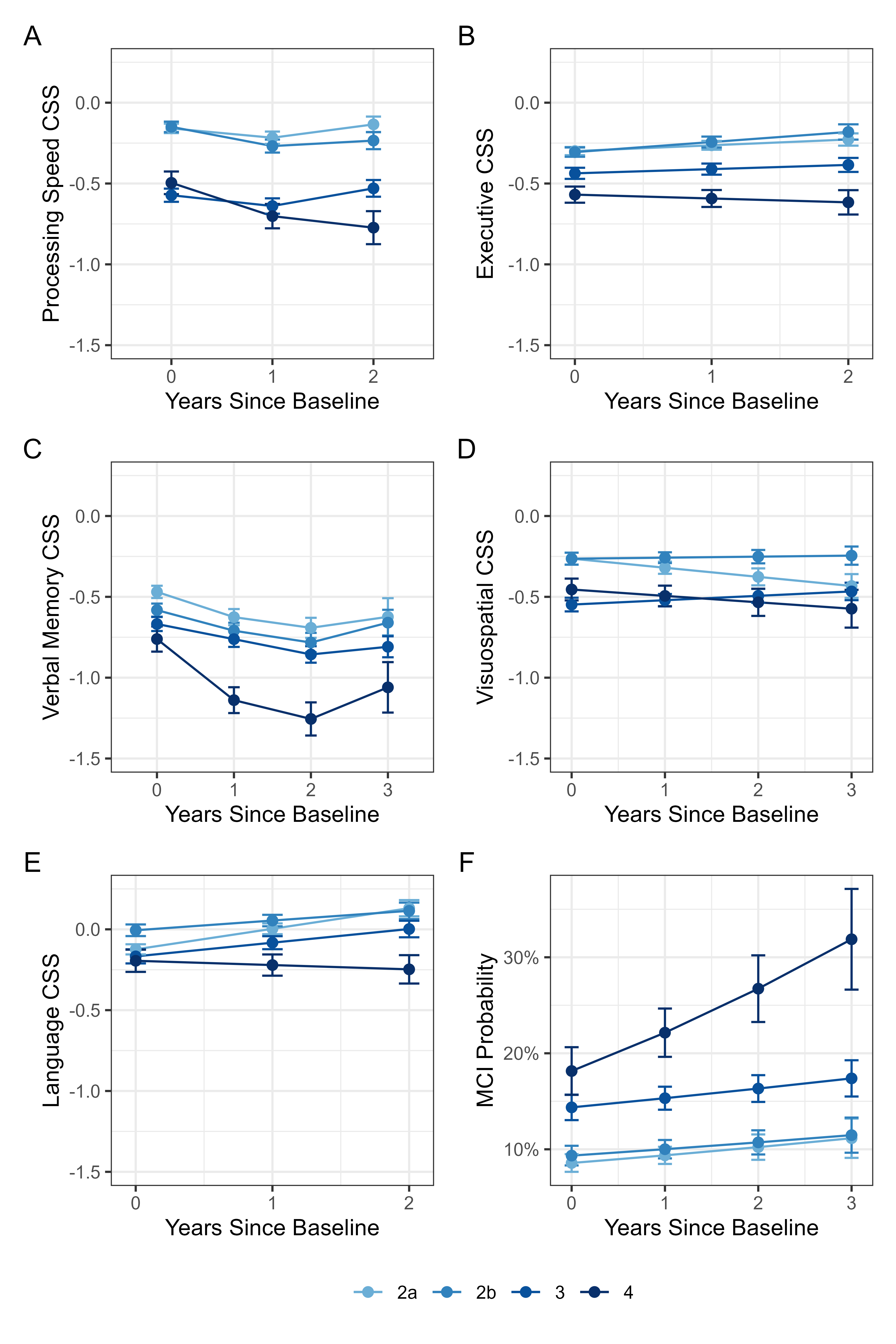

Panels show adjusted trajectories across cognitive domains and MCI among NSD+ participants classified by baseline NSD-ISS stage. Corresponding longitudinal stage tests are summarized in Supplementary Table S10. Because cognition contributes to NSD-ISS stage assignment, these analyses are descriptive and are not used as primary evidence.

**PPMI Study Committees, Cores, and Collaborators**
*June 2025*

**PPMI EXECUTIVE STEERING COMMITTEE**

Kenneth Marek, MD1 (Principal Investigator); Tanya Simuni, MD2 ; Andrew Siderowf, MD3 ; Caroline Tanner, MD4 ; Thomas F Tropea, DO1 ; Tatiana Foroud, PhD5 ; Lana Chahine, MD6 ; Brit Mollenhauer, MD7 ; Kalpana Merchant, MD2 ; Douglas Galasko, MD8 ; Christopher Coffey, PhD9 ; Kathleen Poston, MD10; Roseanne Dobkin, PhD11; Ethan Brown, MD4 ; Roy Alcalay, MD12; Dan Weintraub, MD3 ; Emily Flagg, BA1 ; Kimberly Fabrizio, BA1

**PPMI STEERING COMMITTEE**

Susan Bressman, MD13; Cornelis Blauwendraat, PhD14; Paola Casalin, PhD15; Sonya Dumanis, PhD14; Raymond James, RN16; Karl Kieburtz, MD17; Sneha Mantri, MS18; Werner Poewe, MD19; Michael Schwarzschild, MD20; John Seibyl1 , MD; David Standaert, PhD21; Duygu Tosun-Turgut, PhD4

**MICHAEL J. FOX FOUNDATION** Sohini Chowdhury, MA22; Jamie Eberling, PhD22; Mark Frasier, PhD22; Leslie Kirsch, EdD22; Katie Kopil, PhD22; Maggie Kuhl, BA22; Alyssa O’Grady, BA22; Todd Sherer, PhD22; Tawny Willson, MBS22

**PPMI STUDY CORES**

Project Management Core: Emily Flagg, BA1 Site Management Core: Tanya Simuni, MD2 ; Bridget McMahon, BS1 Data Strategy and Technical Operations: Craig Stanley, PhD1 ; Kim Fabrizio, BA1 Data Management Core: Dixie Ecklund, MBA9 , MSN; Christine Kohnen, PhD9 Screening Core: Tatiana Foroud, PhD5 ; Laura Heathers, BA5 ; Christopher Hobbick, BSCE5 ; Gena Antonopoulos, BSN5 Imaging Core: John Seibyl, MD1 ; Kathleen Poston, MD10 Statistics Core: Christopher Coffey, PhD9 ; Chelsea Caspell-Garcia, MS9 ; Michael Brumm, MS9 Bioinformatics Core: Arthur Toga, PhD23; Karen Crawford, MLIS23 Biorepository Core: Tatiana Foroud, PhD5 ; Jan Hamer, BS5 Biologics Review Committee: Brit Mollenhauer, MD7 ; Doug Galasko, MD8 ; Kalpana Merchant, MD2 Genetics Core: Andrew Singleton, PhD24 Pathology Core: Tatiana Foroud, PhD5 ; Dirk Keene, MD5 Found: Caroline Tanner, MD4 ; Ethan Brown, MD4 PPMI Online: Carlie Tanner, MD4 ; Ethan Brown, MD4 ; Lana Chahine, MD6 ; Roseann Dobkin, PhD11; Monica Korell, MPH4

**PPMI SITE INVESTIGATORS**

Neha Prakash MD1; Tanya Simuni, MD2; Nabila Dahodwala MD3;Caroline Tanner, MD4 ; Lana Chahine MD6 ;Brit Mollenhauer MD7; Sebastian Schade MD7 ; Douglas Galasko, MD 8; Anat Mirelman PhD12; Roy Alcalay MD12;Katherine Leaver MD13; Marie Saint-Hilaire MD16; Ruth Schneider MD17; Christopher Tarolli MD17; Werner Poewe, MD19; Aleksandar Videnovic MD20; David Standaert PhD21; Marissa Dean, MD21; Sonja Jonsdottir PhD25; Rejko Krueger MD25; Claire Pauly PhD25; Stewart Factor DO26; Penelope Hogarth MD26; Robert Hauser MD28; Amy Amara PhD29; Michelle Fullard MD29; Cyrus Zabetian MD30; Hubert Fernandez MD31; Kathrin Brockmann MD32; Isabel Wurster PhD32; Yen Tai PhD33; Paolo Barone PhD34; Marina Picillo MD34; Stuart Isaacson MD35; Alberto Espay MD36; Eduardo Tolosa PhD37; Javier Ruiz Martinez PhD38; Leonidas Stefanis PhD39; Kelvin Chou MD40; Lorraine Kalia MD41; Connie Marras PhD41; David Grimes MD42; Tiago Mestre PhD42; Rajesh Pahwa MD43; Mark Lew MD44; Holly Shill MD45; Shyamal Mehta MD46; Giulietta Riboldi MD47; Nikolaus McFarland PhD48; Ron Postuma MD49; Zoltan Mari MD50; David Ledingham MD51; Nicola Pavese PhD51; Michele Hu PhD52; Norbert Brueggemann MD53, ; Christine Klein MD53; Bastiaan Bloem PhD54; Cristina Simonet PhD55; Alastair Noyce PhD55; Anette Janzen PhD56; David Pedrosa MD56; Wolfgang Oertel PhD56; Njideka Okubadejo MD57David Shprecher DO58; Arjun Tarakad MD59; Emile Moukheiber MD60

**PPMI SITE COORDINATORS** Joy Antala¹; Carla Aranda²; Karen Williams²; Sophia Melton²; Karina Benson²; Ashwini Ramachandran³; Danielle Potts³; Grace LaMoure³; Ritikha Vengadesh3; Ryan Manzler³; Jaime Heller⁴; Primi Ranola⁴; Farah Kausar4; Sherri Mosovsky⁶; Diana Willeke⁷; Elizabeth KalinkaraGomez⁷; Janelle Rodriguez⁸; Nobuko Kemmotsu8; May Eshel¹²; Deborah Raymond¹³; Abigail Desrosiers16; Raymond James¹⁶; Lauren Jackson¹⁷; Iris Egner¹⁹; Wesley Schlett²⁰; Courtney Blair²¹; Lauren Ruffrage²¹; Berenice Sevilla²⁵; Barbara Sommerfeld²⁶; Dustin Le²⁷; Erica Botting²⁸; Gabriella Mazur²⁸; Daniele Derlein²⁹; Evan Doll²⁹; Ying Liu²⁹; Ciera Cobb³⁰; Olivia Masiewicz³⁰; Jennifer Mule³¹; Michael Morsillo³¹; Ella Hilt³²; Aldazier Jakiran33; Dominga Valentino34; Lisbeth Pennente35; Bobbie Stubbeman³⁶; Alicia Garrido³⁷; Valeria Ravasi³⁷; Ioana Croitoru38; Christos Koros³⁹; Nikolas Papagiannakis³⁹; Frank Ferrari⁴⁰; Mengyu Zheng⁴¹; Shawna Reddie⁴²; Alicia Alejandra⁴³; Andrea Gray⁴³; Alejandra Valenzuela44; Caitlin Goodman 45; Sara Dresler⁴⁶; Neil Santos46; Fahrial Esha⁴⁷; Kyle Rizer⁴⁸; Nadine Zablith⁴⁹; Liliana Dumitrescu⁵⁰; Debra Galley⁵¹; Victoria Kate Foster⁵¹; Jamil Razzaque⁵²; Madita Grümmer⁵³; Yara Krasowski⁵⁴; Natalie Donkor⁵⁵; Elisabeth Sittig⁵⁶; Oluwadamilola Ojo⁵⁷; Kelly Clark⁵⁸; Rory Mahabir⁵⁹; Kori Ribb⁶⁰; Shamera Willoughby⁶⁰ June 2025

**INSTITUTIONS AND AFFILIATIONS**

Institute for Neurodegenerative Disorders; New Haven, CT, USA 2. Northwestern University; Evanston, IL, USA 3. University of Pennsylvania; Philadelphia, PA, USA 4. University of California, San Francisco; San Francisco, CA, USA 5. Indiana University; Indianapolis, IN, USA 6. University of Pittsburgh; Pittsburgh, PA, USA 7. Paracelsus-Elena Klinik; Kassel, Germany 8. University of California, San Diego, San Diego, CA, USA 9. University of Iowa; Iowa City, IA, USA 10. Stanford University; Stanford, CA, USA 11. Rutgers University; New Brunswick, NJ, USA 12. Tel Aviv Sourasky Medical Center; Tel Aviv, Israel 13. Mount Sinai Beth Israel; New York, NY, USA 14. Coalition for Aligning Science; Chevy Chase, MD, USA 15. BioRep; Milan, Italy 16. Boston University School of Medicine; Boston, MA, USA 17. University of Rochester; Rochester, NY, USA 18. Duke University; Durham, NC, USA 19. University of Innsbruck; Innsbruck, Austria 20. Massachusetts General Hospital; Boston, MA, USA 21. University of Alabama at Birmingham; Birmingham, AL, USA 22. The Michael J. Fox Foundation for Parkinson’s Research; New York, NY, USA 23. Laboratory of Neuroimaging (LONI), USC; Los Angeles, CA, USA 24. National Institute on Aging, NIH; Bethesda, MD, USA 25. University of Luxembourg; Esch-sur-Alzette, Luxembourg 26. Emory University; Atlanta, GA, USA 27. Oregon Health and Science University; Portland, OR, USA 28. University of South Florida; Tampa, FL, USA 29. University of Colorado; Aurora, CO, USA 30. VA Puget Sound Health System; Seattle, WA, USA 31. Cleveland Clinic; Cleveland, OH, USA 32. University of Tubingen; Tubingen, Germany 33. Imperial College of London; London, UK 34. University of Salerno; Salerno, Italy 35. Parkinson’s Disease and Movement Disorders Center; Boca Raton, FL, USA 36. University of Cincinnati; Cincinnati, OH, USA 37. Hospital Clinic of Barcelona; Barcelona, Spain 38. Hospital Universitario Donostia; San Sebastian, Spain 39. University of Athens; Athens, Greece 40. University of Michigan; Ann Arbor, MI, USA 41. Toronto Western Hospital; Toronto, Canada 42. The Ottawa Hospital; Ottawa, Canada 43. University of Kansas Medical Center; Kansas City, KS, USA 44. Keck School of Medicine of the University of Southern California; Los Angeles, CA, USA 45. Barrow Neurological Institute; Phoenix, AZ, USA 46. Mayo Clinic Arizona; Scottsdale, AZ, USA 47. NYU Langone Medical Center; New York, NY, USA 48. University of Florida; Gainesville, FL, USA 49. Montreal Neurological Institute and Hospital/McGill; Montreal, QC, Canada 50. Cleveland Clinic-Las Vegas Lou Ruvo Center for Brain Health; Las Vegas, NV, USA 51. Clinical Ageing Research Unit; Newcastle, UK 52. John Radcliffe Hospital Oxford and Oxford University; Oxford, UK 53. University of Luebeck; Luebeck, Germany 54. Radboud University; Nijmegen, Netherlands 55. Queen Mary University of London; London, UK 56. Philipps-University Marburg; Marburg, Germany 57. University of Lagos; Lagos, Nigeria 58. Banner Sun Health Research Institute; Sun City, AZ, USA 59. Baylor College of Medicine; Houston, TX, USA 60. Johns Hopkins University; Baltimore, MD, USA
